# *BIN3* rs2280104 T allele is associated with excessive daytime sleepiness and altered network topology in Parkinson’s disease

**DOI:** 10.1101/2023.07.17.23292760

**Authors:** Zhichun Chen, Bin Wu, Guanglu Li, Liche Zhou, Lina Zhang, Jun Liu

## Abstract

**Background:** Excessive daytime sleepiness (EDS) is one of the most common non-motor symptoms in Parkinson’s disease (PD). Previous studies showed that PD patients with EDS exhibited more severe motor and non-motor symptoms. Our recent studies revealed that *BIN3* rs2280104 was negatively associated with scores of Epworth Sleepiness Scale (ESS) in PD patients. The objective of this study is to examine whether *BIN3* rs2280104 shapes brain networks of PD patients and whether network metrics associated with *BIN3* rs2280104 mediate the effects of *BIN3* rs2280104 on EDS.

**Methods:** PD patients (n = 144) receiving functional magnetic resonance imaging in Parkinson’s Progression Markers Initiative (PPMI) database were investigated. The clinical manifestations and graphical metrics of structural and functional network were compared among different genotype groups of *BIN3* rs2280104. The mediation analysis was used to explore the causal associations between network metrics modified by *BIN3* rs2280104 and EDS of PD patients.

**Results:** ESS scores were associated with more severe motor and non-motor symptoms. *BIN3* rs2280104 T allele was negatively associated with ESS scores in PD patients. Additionally, *BIN3* rs2280104 significantly shaped structural and functional network metrics of PD patients. The nodal Cp of left superior temporal pole in functional network and the degree centrality of left calcarine in structural network were negatively associated with ESS scores, however, only the degree centrality of left calcarine in structural network mediated the effects of *BIN3* rs2280104 on EDS of PD patients.

**Conclusions:** To summarize, *BIN3* rs2280104 is significantly associated with EDS and network topology of PD patients. Additionally, the degree centrality of left calcarine in structural network mediated the effects of *BIN3* rs2280104 on EDS. Future studies were required to identify the molecular mechanisms underlying the effects of *BIN3* rs2280104 on EDS and brain network metrics of PD patients.

## Introduction

Parkinson’s disease (PD) is the most common movement disorder characterized by specific motor symptoms, such as static tremor, bradykinesia, rigidity, and postural instability ^1^, however, non-motor symptoms, such as sleep disturbances and autonomic dysfunction, were also very frequent in PD ^1, 2^. There is increasing evidence that the clinical features of PD patients were heterogeneous and complex, which poses a huge challenge to the clinical diagnosis and treatment of PD patients ^3–6^. Therefore, it is extremely important to decipher the neural mechanisms underlying the clinical heterogeneity of PD patients. According to previous literature, the heterogeneity of clinical features in PD patients starts in the prodromal phase ^3^ and prodromal PD can be classified into subtypes with different clinical manifestations, pathomechanisms, imaging markers, and the onset and progression of motor and non-motor symptoms ^3^. It has been proposed that the heterogeneity of clinical manifestations was associated with age ^7, 8^, sex ^7, 8^, environmental factors ^3, 9, 10^, genetic variations ^11–14^, amyloid-β deposits ^15^, tau deposition ^15, 16^, variable spread of synucleinopathy ^3, 15^ and degenerative thresholds ^3, 17^. Accumulated evidence suggests that genetic variations modulate age at onset ^13, 18^, shape motor progression and survival of patients ^19–22^, and affect frequency of diverse non-motor symptoms of PD patients ^12, 23, 24^. Particularly, numerous mutated genes, such as *GBA* ^12, 22, 24^, *LRRK2* ^25–27^, *PRKN* ^28, 29^, *PINK1* ^14, 29^, and *SNCA* ^27, 30^ have been demonstrated to modify the clinical heterogeneity of familial PD patients. In idiopathic PD, a number of genetic variants are found to be associated with the heterogeneity of clinical manifestations ^31–34^. Yoshino et al. (2022) identified 13 novel PRKN variants and revealed that patients with different PRKN variants showed heterogenous clinical features dependent on the number of alternate alleles ^34^. In addition, it has been shown that patients carrying *GBA* variants experienced earlier age at onset and exhibited more severe motor and nonmotor symptoms compared to patients without *GBA* variants ^33, 35^. According to previous studies, the motor function of idiopathic PD patients were associated with variants in *SNCA* ^36, 37^, *BST1* ^38^, *ATP8B2* ^39^, *PARK16* ^40^, and *APOE* ^41^. Additionally, variants of *APOE* ^31, 32^, *SNCA* ^42^, *TMEM106B* ^43^, *COMT* ^44^, *MAPT* ^23, 44^, *PICALM* ^45^ have been found to be associated with variability of cognitive function in idiopathic PD patients. Nevertheless, the associations between genetic variations and clinical features in sporadic PD remain largely elusive and deserved to be further explored.

Excessive daytime sleepiness (EDS) was one of the most common non-motor symptoms in PD ^46–49^. According to a recent meta-analysis, the prevalence of EDS was 35% in PD and EDS was associated with worse motor and autonomic dysfunction, reduced autonomy, and higher burden of neuropsychiatric symptoms ^49^. Additionally, EDS was also associated with increased frequency falls ^50^, sarcopenia ^50^, development of swallowing impairment ^51^, and poorer quality of life (QoL) ^52, 53^. In prodromal PD, EDS was recognized to be a significant predictor of future neurodegeneration ^54^. According to previous literature, a multitude of risk factors have been found to be associated with EDS, including male sex ^55, 56^, age ^55, 57^, cognitive impairment ^55, 56^, worse sleep quality ^55–57^, autonomic dysfunction ^58^, depression ^58^, rapid eye movement sleep behavior disorder (RBD) ^55, 58^, dopaminergic therapy ^48, 56, 57^, use of antihypertensives ^56^, and worse QoL ^55^. In addition, genetic variations were also associated with EDS. For example, Adam et al. (2022) reported that narcolepsy Human Leukocyte Antigen (*HLA*) risk allele DQB1*06:02 was significantly associated with EDS and DQB1*06:02-positive PD patients usually and inappropriately fell asleep during daytime activities requiring sustained alertness ^59^. Recently, we found that a PD risk-associated genetic variant, *BIN3* rs2280104 (C > T, C is the major allele and T is the minor allele) ^60, 61^ was significantly associated with the scores of Epworth Sleepiness Scale (ESS) in PD patients ^62^. *BIN3* rs2280104 T allele was associated with higher prevalence of PD compared to C alllele ^60, 61^. Based on Genotype-Tissue Expression (eQTL) database (https://www.gtexportal.org/home/), *BIN3* rs2280104 was also associated with differential expressions and splicing of multiple genes in brain tissue ^62^. Therefore, *BIN3* rs2280104 is a key genetic variant contributing to disease susceptibility, brain gene expressions, and EDS of PD patients.

Although EDS was an essential non-motor symptom of PD, the neural mechanisms underlying the development of EDS remain largely unknown. Recently, we found brain structural and functional network metrics were significantly associated with motor and non-motor symptoms of PD patients ^7, 23, 62^. In addition, we also revealed that small-worldness properties of white matter network mediated the effects of *MAPT* rs17649553 on verbal memory of PD patients ^23^. Furthermore, small-world topology in gray matter covariance network specifically mediated the effects of *OGFOD2/CCDC62* rs11060180 on motor function of PD patients ^62^. Therefore, PD-associated risk genes may modify the motor and non-motor symptoms by shaping brain structural and functional networks ^23, 62^. Based on these findings, we hypothesized that *BIN3* rs2280104 may also affect brain network metrics and network metrics associated with *BIN3* rs2280104 may mediate the effects of *BIN3* rs2280104 on EDS. In this study, we aim to investigate whether *BIN3* rs2280104 modulates the structural and functional brain networks of PD patients and whether network metrics shaped by *BIN3* rs2280104 mediate its associations with EDS. Particularly, our objectives include: (i) examine whether *BIN3* rs2280104 T allele is associated with EDS in PD patients using association analysis and χ^2^ test; (ii) compare the differences of clinical features between EDS+ and EDS-patients; (iii) investigate whether structural and functional network metrics are statistically different between CC carriers and T-carriers (CT and TT carriers); and (iv) evaluate whether the structural and functional network metrics mediate the effects of *BIN3* rs2280104 on EDS of PD patients.

## Materials and Methods

### Participants

This study is based on data acquired within Parkinson’s Progression Markers Initiative (PPMI) study, a population-based cohort study. The study was approved by Institutional Review Board of all the participating sites. All participants signed informed consent, which can be obtained from the site investigators. The criteria for PD patients included: (i) The patients were aged > 30 years old; (ii) The patients were diagnosed as PD according to the Movement Disorders Society (MDS) Clinical Diagnostic Criteria for PD; (iii) The patients had a disease duration ≤2 years; (iv) The patients were not on dopamine replacement therapy; (v) The patients acquired 3D T1-weighted MPRAGE imaging, resting-state fMRI imaging, and diffusion tensor imaging (DTI) during the same period. The exclusion criteria for PD patients were shown below: (i) The patients had major abnormalities in the standard MRI; (ii) The patients were diagnosed with dementia with PD or dementia with Lewy bodies according to the MDS criteria; (iii) The patients were treated with neuroleptics, metoclopramide, α-methyldopa, methylphenidate, reserpine, amphetamine derivative, or hypnotic medications; (iv) The patients carried genetic mutations of familial PD or were from the genetic PPMI cohort and prodromal cohort. The motor manifestations of PD patients were assessed with Hoehn and Yahr (H&Y) stages, tremor scores, total rigidity scores, and the MDS Unified Parkinson’s Disease Rating Scale (MDS-UPDRS). The non-motor symptoms were evaluated with Epworth Sleepiness Scale (ESS), Scale for Outcomes in Parkinson’s Disease-Autonomic (SCOPA-AUT), 15-item Geriatric Depression Scale (GDS), REM Sleep Behavior Disorder Screening Questionnaire (RBDSQ), Symbol Digit Modalities Test (SDMT), Montreal Cognitive Assessment (MoCA), Benton Judgment of Line Orientation (BJLOT) test, Letter Number Sequencing (LNS) test, and Semantic Fluency Test (SFT). The patients also received [^123^I] FP-CIT SPECT images, which were analyzed following the technical operations manual (http://ppmi-info.org/). The striatal binding ratios (SBRs) for caudate, putamen, and striatum were extracted from SPECT scans. The SBRs were computed as (target region/reference region)-1, where the reference region was the occipital lobe. To assess the effects of EDS on clinical assessments and brain networks, PD patients were divided into EDS+ group (ESS > 10) and EDS-group (ESS ≤ 10). The ESS is a self-administered questionnaire developed in 1990 by Australian doctor Murray Johns. The scores of ESS range between 0 and 24. Scores between 0 to 10 reflect normal levels of daytime sleepiness while scores over 10 are considered to have EDS. The demographic data of participants included for each group were shown in Table S1, showing comparable demographic features between EDS-patients and EDS+ patients. To explore the associations between *BIN3* rs2280104 (C > T) and EDS or brain network metrics, the patients also received whole-exome sequencing of blood DNA samples and obtained the genotypes of *BIN3* rs2280104 for each participant. The descriptive statistics of clinical characteristics in different genotype groups (CC carriers, CT carriers, TT carriers) of *BIN3* rs2280104 were shown in Table 1.

**Table 1.** The demographic and clinical data for different genotype groups of *BIN3* rs2280104. The data were shown as the mean ± standard deviation (SD). The motor function examination was assessed in ON state. Unpaired t-test (CC carriers *vs* T-carriers) and one-way ANOVA followed by Tukey’s post hoc test (CC carriers *vs* CT carrier *vs* TT carriers) were used to compare the clinical variables among different genotype groups of *BIN3* rs2280104. Abbreviations: HY, Hoehn & Yahr stage; UPDRS-III, Unified Parkinson’ s Disease Rating Scale Part III; RBDSQ, REM Sleep Behavior Disorder Screening Questionnaire; SCOPA-AUT, Scale for Outcomes in Parkinson’s Disease-Autonomic; SDMT, Symbol Digit Modalities Test; LNS, Letter Number Sequencing; BJLOT, Benton Judgement of Line Orientation; MoCA, Montreal Cognitive Assessment.

| Clinical variable | CC carriers<br>(n = 53) | T-carriers<br>(n = 93) | CT carriers<br>(n = 72) | TT carriers<br>(n = 21) | CC carriers<br>vs T-carriers | CC carriers vs<br>CT carriers | CC carriers vs<br>TT carriers |
| --- | --- | --- | --- | --- | --- | --- | --- |
| Age, years | 61.16 ± 8.74 | 61.29 ± 9.52 | 61.16 ± 9.17 | 61.71 ± 10.86 | $p > 0.05$ | $p > 0.05$ | $p > 0.05$ |
| Sex (Male/Female) | 34/19 | 60/33 | 47/25 | 13/8 | $p > 0.05$ | $p > 0.05$ | $p > 0.05$ |
| Education, years | 15.15 ± 2.88 | 15.33 ± 3.06 | 15.35 ± 2.92 | 15.29 ± 3.59 | $p > 0.05$ | $p > 0.05$ | $p > 0.05$ |
| Disease duration, years | 2.43 ± 2.86 | 1.82 ± 1.58 | 1.92 ± 1.68 | 1.45 ± 1.17 | $p > 0.05$ | $p > 0.05$ | $p > 0.05$ |
| HY | 1.65 ± 0.56 | 1.67 ± 0.47 | 1.62 ± 0.49 | 1.86 ± 0.36 | $p > 0.05$ | $p > 0.05$ | $p > 0.05$ |
| Tremor | 3.73 ± 3.02 | 3.87 ± 3.40 | 3.94 ± 3.43 | 3.62 ± 3.38 | $p > 0.05$ | $p > 0.05$ | $p > 0.05$ |
| Rigidity | 4.35 ± 3.03 | 4.25 ± 2.71 | 4.21 ± 2.81 | 4.38 ± 2.38 | $p > 0.05$ | $p > 0.05$ | $p > 0.05$ |
| UPDRS-III | 20.69 ± 11.39 | 20.95 ± 8.58 | 20.73 ± 8.47 | 21.67 ± 9.13 | $p > 0.05$ | $p > 0.05$ | $p > 0.05$ |
| ESS | **7.09 ± 4.21** | **5.94 ± 3.67** | **6.26 ± 3.86** | **4.81 ± 2.70** | **$p = 0.0843$** | **$p > 0.05$** | **$p = 0.0593$** |
| RBDSQ | 4.13 ± 2.54 | 4.32 ± 2.74 | 4.51 ± 2.93 | 3.67 ± 1.91 | $p > 0.05$ | $p > 0.05$ | $p > 0.05$ |
| SCOPA-AUT | 10.27 ± 6.17 | 9.75 ± 5.33 | 10.23 ± 5.40 | 8.14 ± 4.85 | $p > 0.05$ | $p > 0.05$ | $p > 0.05$ |
| LNS | 11.08 ± 2.94 | 10.25 ± 2.80 | 10.26 ± 2.76 | 10.19 ± 2.99 | $p > 0.05$ | $p > 0.05$ | $p > 0.05$ |
| BJLOT | 12.92 ± 1.89 | 12.63 ± 2.30 | 12.69 ± 2.24 | 12.43 ± 2.54 | $p > 0.05$ | $p > 0.05$ | $p > 0.05$ |
| SDMT | 40.08 ± 12.07 | 40.66 ± 10.23 | 40.63 ± 10.81 | 40.76 ± 8.17 | $p > 0.05$ | $p > 0.05$ | $p > 0.05$ |
| MoCA | 26.64 ± 3.16 | 27.17 ± 2.38 | 27.03 ± 2.32 | 27.67 ± 2.54 | $p > 0.05$ | $p > 0.05$ | $p > 0.05$ |

### Image acquisition

The three-dimensional T1-weighted MRI images were acquired on 3T Siemens (TIM Trio and Verio) scanners (Erlangen, Germany) using a magnetisation-prepared rapidacquisition gradient echo sequence. The parameters for T1 images were shown as follows: TR = 2300 ms, TE = 2.98 ms, Voxel size = 1 mm^3^, Slice thickness =1.2 mm, twofold acceleration, sagittal-oblique angulation. The diffusion tensor imaging was performed with the parameters below: TR= 8,400-8,800 ms, TE = 88 ms, Voxel size = 2 mm^3^, Slice thickness = 2 mm, 64 directions, b = 1000 s/mm^2^. The resting-state images were acquired using a T2*-weighted echo planar imaging (EPI) sequence with the parameters as follows: TR = 2400 ms, TE = 25 ms, Voxel size = 3.3 mm^3^, Slice number = 40, Slice thickness = 3.3 mm, Flip angle = 90°, Acquisition time = 8 min.

### Imaging preprocessing

Diffusion MRI images were preprocessed using FMRIB Software Library toolbox (FSL, https://fsl.fmrib.ox.ac.uk/fsl/fslwiki). In brief, the diffusion images were corrected for subject motion, eddy current distortions, and susceptibility artifacts due to the magnetic field inhomogeneity. The DTI metrics including fractional anisotropy (FA), mean diffusivity (MD), axial diffusivity, and radial diffusivity maps were derived. Then, the individual images were reconstructed in the standard MNI space for comparisons across participants.

The preprocessing of resting-state images was performed using SPM12 (http://www.fil.ion.ucl.ac.uk/spm/software/spm12/) and GRETNA software (https://www.nitrc.org/projects/gretna/). As shown by our recent studies, the preprocessing steps included regular procedures for slice-timing correction, spatial normalizing using standard EPI template, spatial smoothing (4-mm Gaussian kernel), removal of nuisance signals (head motion profiles, CSF signal, white matter signals, and local and global hardware artifacts), temporal bandpass filtering (0.01–0.1 Hz). The 9 of 83 participants with head motion frame-wise displacement (FD) > 0.5 mm and head rotation > 2° were excluded according to previous studies ^63–66^ due to head movements. Furthermore, the head motion parameters were regressed out to adjust the effect of head motions on the functional images. Finally, we found there were no statistical differences in head motor FD between the EDS+ patients and the EDS-patients or between CC carriers and T-carriers (*p* > 0.05).

### Network construction

A free and open MATLAB toolkit, PANDA (http://www.nitrc.org/projects/panda/) was utilized to perform deterministic fiber tractography to construct the white matter network. The Fiber Assignment by Continuous Tracking (FACT) algorithm was used to calculate whole-brain white matter fibers between each pair of 90 cortical and subcortical nodes in the AAL atlas. The mean FA skeleton was then thresholded to FA > 0.20 and the white matter fiber angle was thresholded to 45°. After the tractography, a white matter network matrix based on fiber number (FN) between two nodes of AAL atlas was created for each individual.

The construction of functional connectivity matrix between individual regions was performed as follows. Firstly, ninety cortical and subcortical nodes using the AAL atlas were defined. Secondly, the pairwise function connectivity was calculated as the Pearson correlation coefficient of the time series of 90 nodes. The Fisher’s r-to-z transformations of correlation coefficients were performed to improve the normality of functional connectivity. Finally, a 90 x 90 functional connectivity matrix were constructed for each participant.

### Graph-based network analysis

The graphical network metrics of structural and functional networks were calculated using GRETNA (https://www.nitrc.org/projects/gretna/) ^67^. To calculate the global and nodal network metrics, a series of network sparsity threshold (0.05 ∼ 0.50 with an interval of 0.05) was set. The area under curve (AUC) of global and nodal network metrics was computed. The global network properties included assortativity, hierarchy, global efficiency, local efficiency, and small-worldness metrics. The small-worldness properties included: clustering coefficient (Cp), characteristic path length (Lp), normalized clustering coefficient (γ), normalized characteristic path length (λ), and small worldness (σ). The nodal network properties included: nodal betweenness centrality, nodal degree centrality, nodal Cp, nodal efficiency, nodal local efficiency, and nodal shortest path length. The definitions for each network metric have been reported by previous studies ^62, 68, 69^.

## Statistical analysis

### Comparison of clinical variables

Unpaired t-test (CC carriers vs T-carriers or EDS-patients *vs* EDS+ patients) and one-way ANOVA followed by Tukey’s post hoc test (CC carriers vs CT carrier vs TT carriers) were used to compare continuous variables. χ^2^ test was used to compare categorical variables. *p* < 0.05 was considered statistically significant.

### Comparison of global network strength

Two sample t-test in Network-Based Statistic (NBS, https://www.nitrc.org/projects/nbs/) software was used to compare the global network strength of structural and functional networks between CC carriers and T-carriers ^70^. *p* < 0.05 after false discovery rate (FDR) correction was considered statistically significant ^71^. The covariates, such as age, sex, years of education, and disease duration, were included during the analysis.

### Comparison of network metrics

For comparisons of global and nodal network metrics, two-way ANOVA test followed by FDR corrections was used. *p* < 0.05 after FDR correction was considered statistically significant. The AUCs of global network metrics between CC carriers and T-carriers were compared using two sample t-test and *p* < 0.05 was considered statistically significant.

### Association analysis

The association analysis between number of T alleles and ESS scores was conducted by Spearman’s correlation method. The univariate correlation analysis between ESS scores and network metrics was performed with Pearson correlation method. The multivariate regression analysis was utilized to analyze the associations between network metrics and ESS scores or between ESS scores and clinical assessments or between *BIN3* rs2280104 and graphical network metrics with age, sex, disease duration, and years of education as covariates. We examined whether multicollinearity occurred during multivariate regression analysis using IBM SPSS Statistics Version 26. *p* < 0.05 was considered statistically significant for associations between number of T alleles and ESS scores or between network metrics and ESS scores or between ESS scores and clinical assessments. FDR-corrected *p* < 0.05 was considered statistically significant for associations between *BIN3* rs2280104 and graphical network metrics.

### Mediation analysis

IBM SPSS Statistics Version 26 was utilized to perform mediation analysis. The independent variable in the mediation model was the genotype of *BIN3* rs2280104. The dependent variable was ESS scores or EDS. The mediators were topological network metrics associated with ESS scores or EDS. We modeled the mediated effects of network metrics on the relationships between ESS scores and genotype of *BIN3* rs2280104 or between EDS and genotype of *BIN3* rs2280104. The age, sex, disease duration, and years of education were included as covariates during the mediation analysis. *p* < 0.05 was considered statistically significant.

## Results

### Allele and genotype frequencies

The allele frequencies (C allele frequency = 0.6088, T allele frequency = 0.3912) for *BIN3* rs2280104 were similar to the results from Allele Frequency Aggregator Project (T allele frequency = 0.3428). The distribution of genotype frequencies from our patients matches Hardy– Weinberg equilibrium (*p* > 0.10). Genotype frequencies distribute similarly with regard to age or sex (*p* > 0.05).

### Group difference of clinical variables

In 146 PD patients, EDS+ patients (n = 17) exhibited higher ESS scores compared to EDS-patients (n = 129). The demographic characteristics of EDS+ patients and EDS-patients were shown and compared in Table S1, indicating age, sex, years of education, and disease duration were comparable between two groups. Compared to EDS-patients, EDS+ patients showed higher rigidity scores (4.11 ± 2.75 *vs* 5.63 ± 3.12, *p* < 0.05; Fig.1A), UPDRS-III scores (20.23 ± 9.37 *vs* 25.81 ± 10.62, *p* < 0.05; Fig.1B), UPDRS total scores (32.00 ± 13.66 *vs* 47.00 ± 18.46, *p* = 0.0001; Fig.1C), RBDSQ scores (4.01 ± 2.54 *vs* 6.12 ± 2.96, *p* < 0.01; Fig.1D), SCOPA-AUT scores (9.42 ± 5.40 *vs* 13.82 ± 5.97, *p* < 0.01; Fig.1E) and lower SDMT scores (41.21 ± 10.73 *vs* 34.65 ± 10.72, *p* < 0.05; Fig.1F). In addition, EDS+ patients also had lower SBRs in right caudate (1.83 ± 0.53 vs 1.57 ± 0.61, *p* = 0.07; Fig.S1A), left caudate (1.82 ± 0.58 *vs* 1.49 ± 0.56, *p* < 0.05; Fig.S1B), right putamen (0.76 ± 0.32 *vs* 0.58 ± 0.27, *p* < 0.05; Fig. S1C), left putamen (0.75 ± 0.34 *vs* 0.58 ± 0.20, *p* = 0.05; Fig. S1D), right striatum (2.59 ± 0.79 *vs* 2.15 ± 0.86, *p* < 0.05; Fig. S1E), left striatum (2.57 ± 0.89 *vs* 2.08 ± 0.71, *p* < 0.05; Fig. S1F), bilateral caudate (1.82 ± 0.52 *vs* 1.53 ± 0.55, *p* < 0.05; Fig. S1G), bilateral putamen (0.76 ± 0.29 *vs* 0.58 ± 0.20, *p* < 0.05; Fig. S1H), and bilateral striatum (1.29 ± 0.38 *vs* 1.06 ± 0.36, *p* < 0.05; Fig. S1I).

**Fig. 1.**
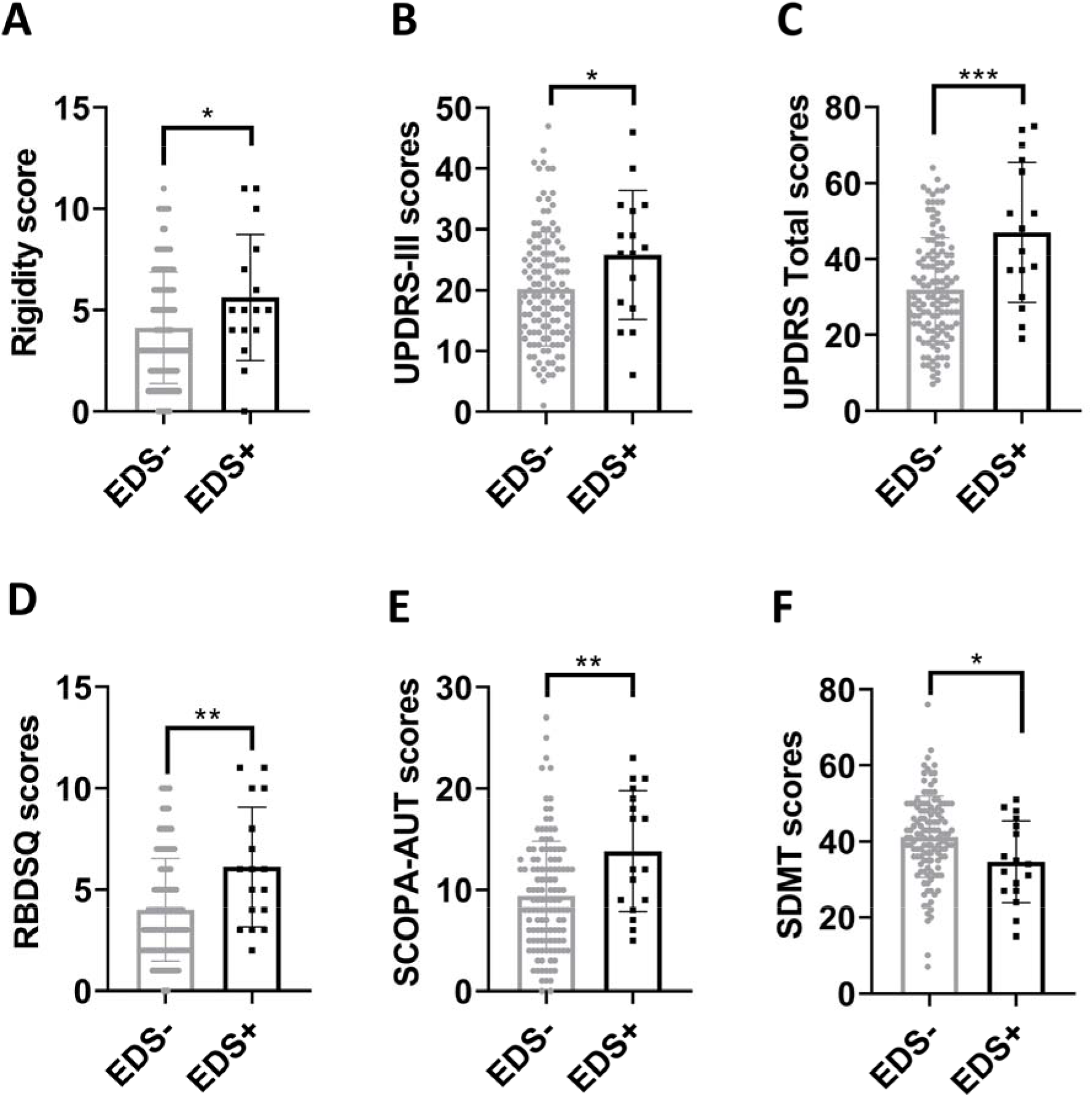
Worse motor and non-motor symptoms in EDS+ patients compared to EDS-patients. (A-F) Group differences of Rigidity scores (A), UPDRS-III scores (B), UPDRS Total scores (C), RBDSQ scores (D), SCOPA-AUT scores (E), and SDMT scores (F). Unpaired t-test was used to compare the clinical variables between EDS-patients and EDS+ patients. *p* < 0.05 was considered statistically significant. \**p* < 0.05, \*\**p* < 0.01, \*\*\**p* < 0.001.

The statistical differences in the clinical variables of different genotype groups of *BIN3* rs2280104 were shown in Table 1. Except ESS scores, T-carriers exhibited comparable levels of clinical variables compared to CC carriers (Table 1).

### The associations between ESS scores and clinical assessments

The associations between ESS scores and clinical assessments of PD patients were shown in Table S2. The ESS scores were positively associated with scores of Rigidity (β = 0.15, *p* = 0.0192), UPDRS-III (β = 0.60, *p* = 0.0042), RBDSQ (β = 0.27, *p* < 0.0001), and SCOPA-AUT (β = 0.59, *p* < 0.0001) in PD patients. The ESS scores were negatively associated with scores of BJLOT (β = –0.11, *p* = 0.0063), SDMT (β = –0.71, *p* = 0.0006), and MoCA (β = –0.18, *p* = 0.0013). In addition, ESS scores were also negatively associated with SBRs of striatum subregions (Table S2).

### The associations between *BIN3* rs2280104 and EDS

In a previous study, we reported a PD-associated risk variant, *BIN3* rs2280104 was associated with ESS scores in 198 PD patients. Consistently, we also observed a significant association between *BIN3* rs2280104 and ESS scores in current study (β = –1.16, *p* < 0.05, n = 146; Fig.2A), which is independent of age, sex, years of education, and disease duration. Compared to CC carriers (7.09 ± 4.21), T-carriers (5.94 ± 3.67) exhibited lower ESS scores approaching borderline significance (*p* = 0.0843; Fig.2B). Additionally, TT carriers (4.81 ± 2.70) showed much lower ESS scores compared to CC carriers (7.09 ± 4.21), close to a marginally significant level (*p* = 0.0593; Fig.2C). Using χ^2^ test, we further demonstrated that EDS was much less frequent in T-carriers (6.45%, CT and TT carriers) than CC carriers (20.75%, *p* < 0.01 and *p* < 0.05; Fig.2D-E).

**Fig. 2.**
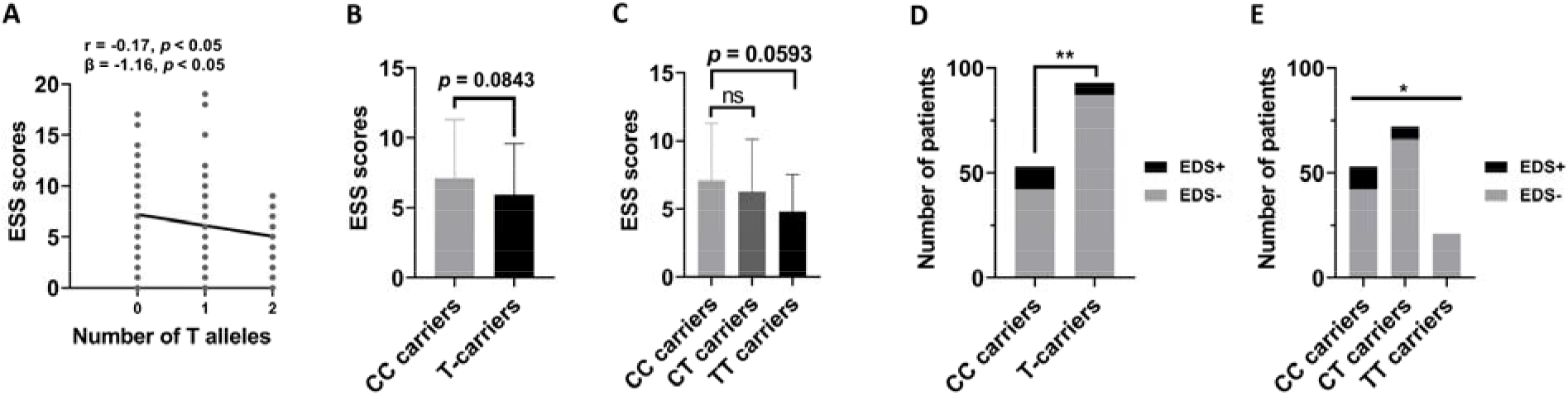
*BIN3* rs2280104 T allele was negatively associated with EDS in PD patients. (A) The number of T alleles was negatively associated with ESS scores (*p* < 0.05 in both Pearson correlation analysis and multivariate regression analysis). (B) T-carriers exhibited lower ESS scores, approaching borderline significance (*p* = 0.0843). (C) TT carriers showed much lower ESS scores compared to CC carriers, which is close to a marginally significant level (*p* = 0.0593). (D) The frequency of EDS was lower in T-carriers compared to CC carriers (*p* < 0.01). (E) The frequency of EDS was lower in TT carriers and CT carriers compared to CC carriers (*p* < 0.05). The association analysis between number of T alleles and ESS scores was conducted by Spearman’s correlation method and multivariate regression analysis with age, sex, disease duration, and years of education as covariates. Unpaired t-test (CC carriers *vs* T-carriers) and one-way ANOVA followed by Tukey’s post hoc test (CC carriers *vs* CT carrier *vs* TT carriers) were used to compare ESS scores among different genotype groups of *BIN3* rs2280104. χ^2^ test was used to compare the distribution of EDS in different genotype groups of *BIN3* rs2280104. \**p* < 0.05, \*\**p* < 0.01.

### Group difference of global connectivity strengths

The global connectivity strengths of structural network were not statistically different between CC carriers and T-carriers (FDR-corrected *p* > 0.05, NBS method, 10,000 permutations). Additionally, the global connectivity strengths of functional network were also not significantly different between CC carriers and T-carriers (FDR-corrected *p* > 0.05, NBS method, 10,000 permutations).

### Group difference of graphical metrics in structural network

The global efficiency of structural network was much higher in T-carriers than that of CC carriers at multiple network sparsity (*p* < 0.01 at network sparsity = 0.15 – 0.5, FDR-corrected; Fig.3A). The small-wordless Lp tended to be lower in T-carriers compared to that of CC carriers (*p =* 0.0528 at network sparsity = 0.15 – 0.5, FDR-corrected; Fig.3B). Consistently, the AUCs of global efficiency were higher in T-carriers while the AUCs of small-worldness Lp were lower in T-carriers (unpaired t-test, *p* < 0.05; Fig.3C-D). The other global network metrics were not statistically different between CC carriers and T-carriers (FDR-corrected *p* > 0.05, data not shown). For nodal network metrics, the betweenness centrality in left middle occipital gyrus and left putamen were lower in T-carriers compared to CC carriers (*p* < 0.0001, FDR-corrected; Fig.3E). The other nodal metrics in structural network were not statistically different between CC carriers and T-carriers (FDR-corrected *p* > 0.05, data not shown).

**Fig. 3.**
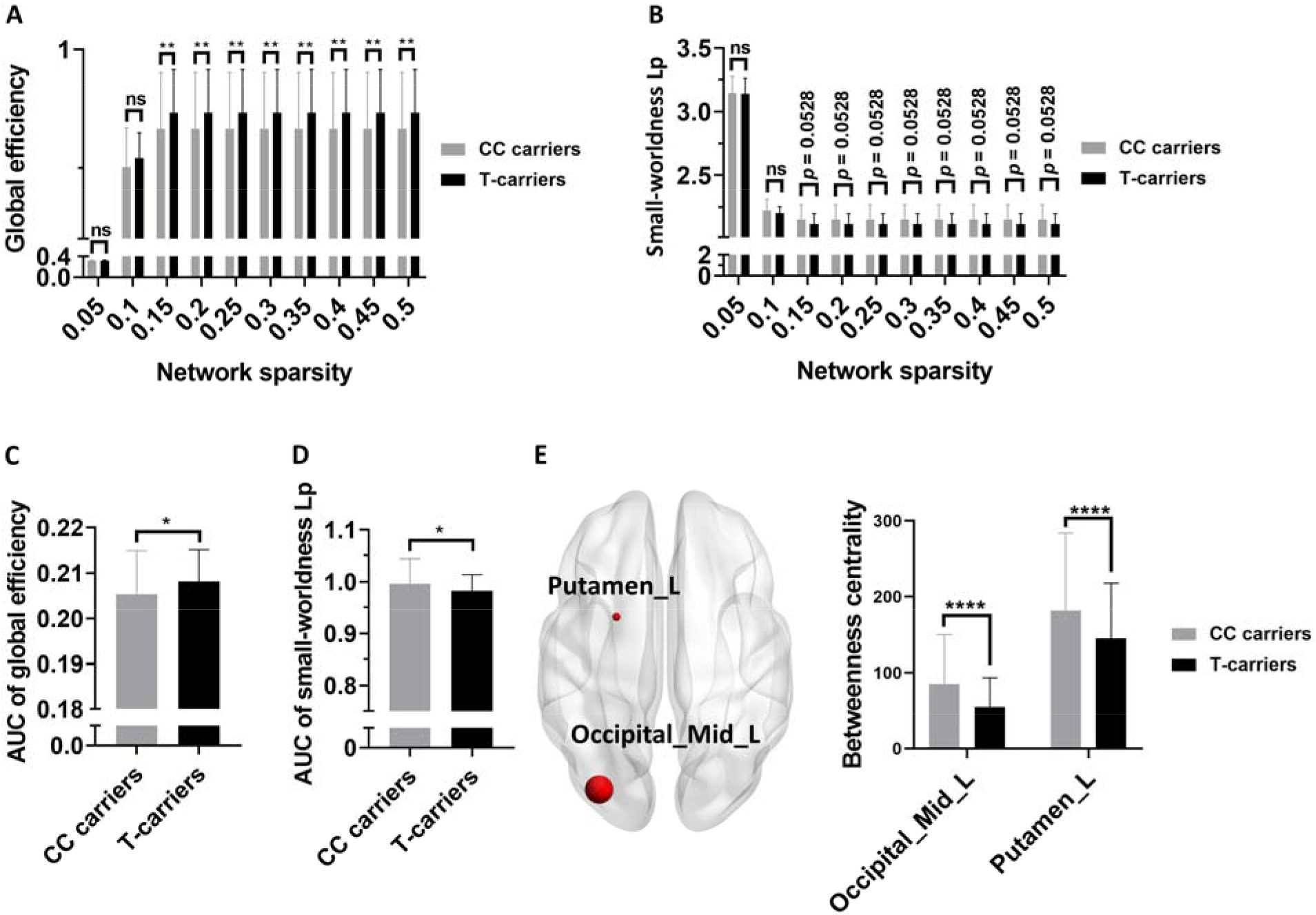
Group differences of structural network metrics between CC carriers and T-carriers. (A-E) Group differences of global efficiency at multiple network sparsity (A), small-worldness Lp at multiple network sparsity (B), AUC of global efficiency (C), AUC of small-worldness Lp (D), nodal betweenness centrality (E) in structural network. Two-way ANOVA test followed by FDR corrections was used for the analysis of results in A, B, and E (*p* < 0.05 after FDR correction was considered statistically significant). The AUCs of global network metrics between CC carriers and T-carriers were compared using unpaired t-test (*p* < 0.05 was considered statistically significant). \**p* < 0.05, \*\**p* < 0.01, \*\*\*\**p* < 0.0001. Abbreviations: Lp, characteristic path length; AUC, Area under curve.

To further evaluate the dose effects of T allele on structural network metrics of PD patients, we compared the differences of structural network metrics among CC carriers, CT carriers, and TT carriers. The global network metrics were not statistically different among different genotype groups of *BIN3* rs2280104 (FDR-corrected *p* > 0.05). For nodal network metrics, CT and TT carriers showed lower betweenness centrality in left middle occipital gyrus, left putamen, and right putamen compared to CC carriers (FDR-corrected *p* < 0.05; Fig. 4A). The degree centrality of left calcarine, left cuneus, and left superior occipital gyrus were higher in CT and TT carriers compared to CC carriers (FDR-corrected *p* < 0.05; Fig. 4B). The nodal Cp of right amygdala, left inferior occipital gyrus, and left angular gyrus were higher in TT carriers compared to CC and CT carriers (FDR-corrected *p* < 0.05; Fig. 4C). The nodal local efficiency of right middle frontal orbital gyrus and right amygdala in CT and TT carriers were much higher than those of CC carriers (FDR-corrected *p* < 0.05; Fig. 4D).

**Fig. 4.**
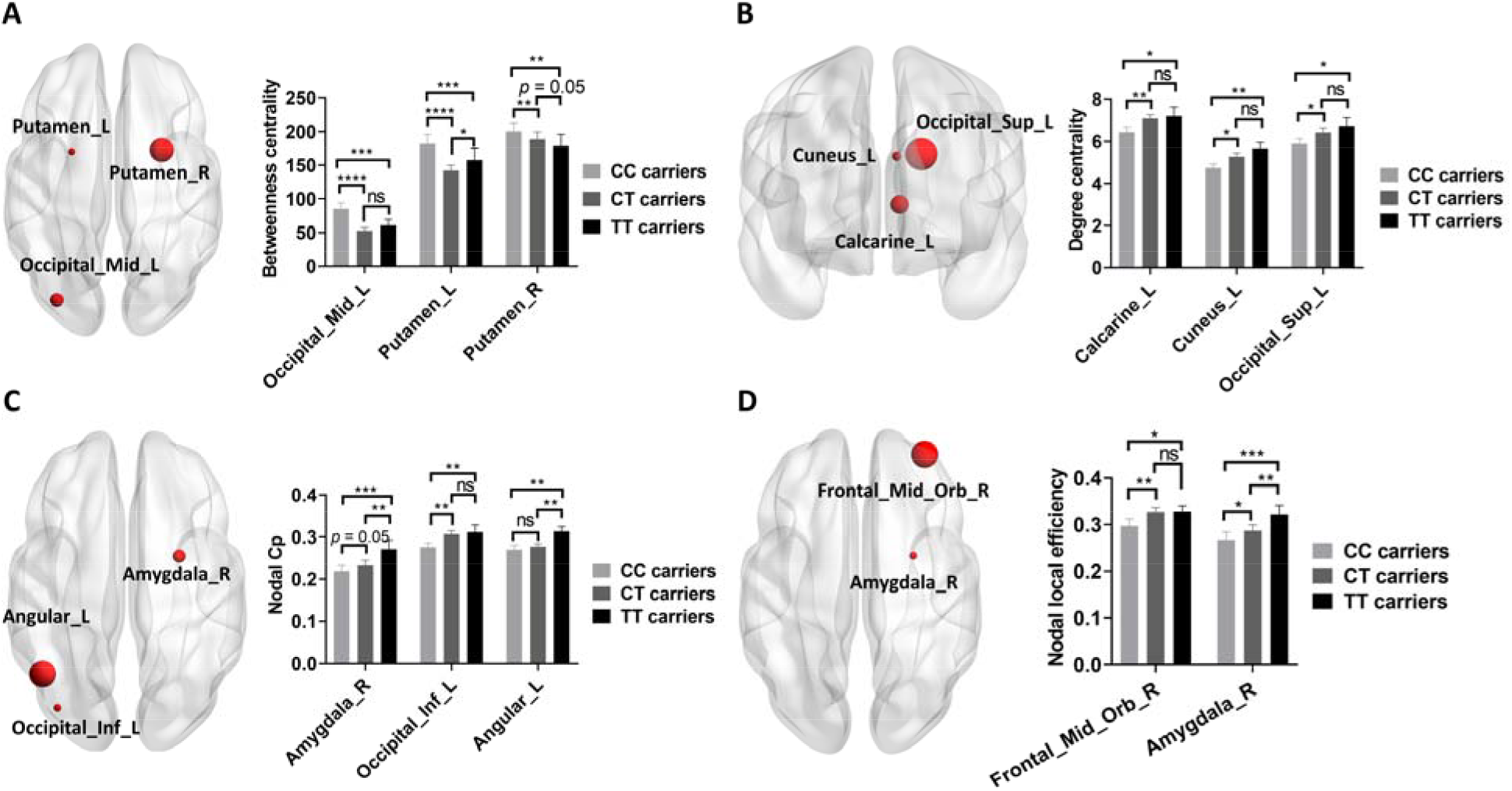
Group differences of structural network metrics among CC carriers, CT carriers, and TT carriers. (A-D) Group differences of nodal betweenness centrality (A), nodal degree centrality (B), nodal Cp (C), and nodal local efficiency (D) in structural network. Two-way ANOVA test followed by FDR corrections was used and *p* < 0.05 after FDR correction was considered statistically significant. \**p* < 0.05, \*\**p* < 0.01, \*\*\**p* < 0.001, \*\*\*\**p* < 0.0001. Abbreviations: Cp, Clustering coefficient.

### Group difference of graphical metrics in functional network

In functional network, the hierarchy (FDR-corrected *p* < 0.05 at sparsity = 0.3 – 0.5; Fig. 5A) and global efficiency (FDR-corrected *p* < 0.05 at sparsity = 0.05 – 0.15; Fig. 5B) were much lower in T-carriers compared to CC carriers. In agreement with these findings, the AUCs of hierarchy and global efficiency were also lower in T-carriers compared to CC carriers (unpaired t-test, *p* < 0.05; Fig.5C-D). For nodal network metrics, the nodal Cp, nodal efficiency, nodal local efficiency of left caudate were lower and nodal shortest path length was higher in T-carriers compared to CC carriers (FDR-corrected *p* < 0.05; Fig.5E-H). The nodal local efficiency of right caudate was also lower in T-carriers, companied by higher nodal shortest path length of right caudate (FDR-corrected *p* < 0.05; Fig.5G-H). In contrast, the nodal Cp and nodal local efficiency of right middle temporal pole were greater in T-carriers compared to CC carriers (FDR-corrected *p* < 0.05; Fig.5E and 5G). Additionally, we also found higher nodal shortest path length of bilateral post cingulate gyrus in T-carriers compared to CC carriers (FDR-corrected *p* < 0.05; Fig.5H).

**Fig. 5.**
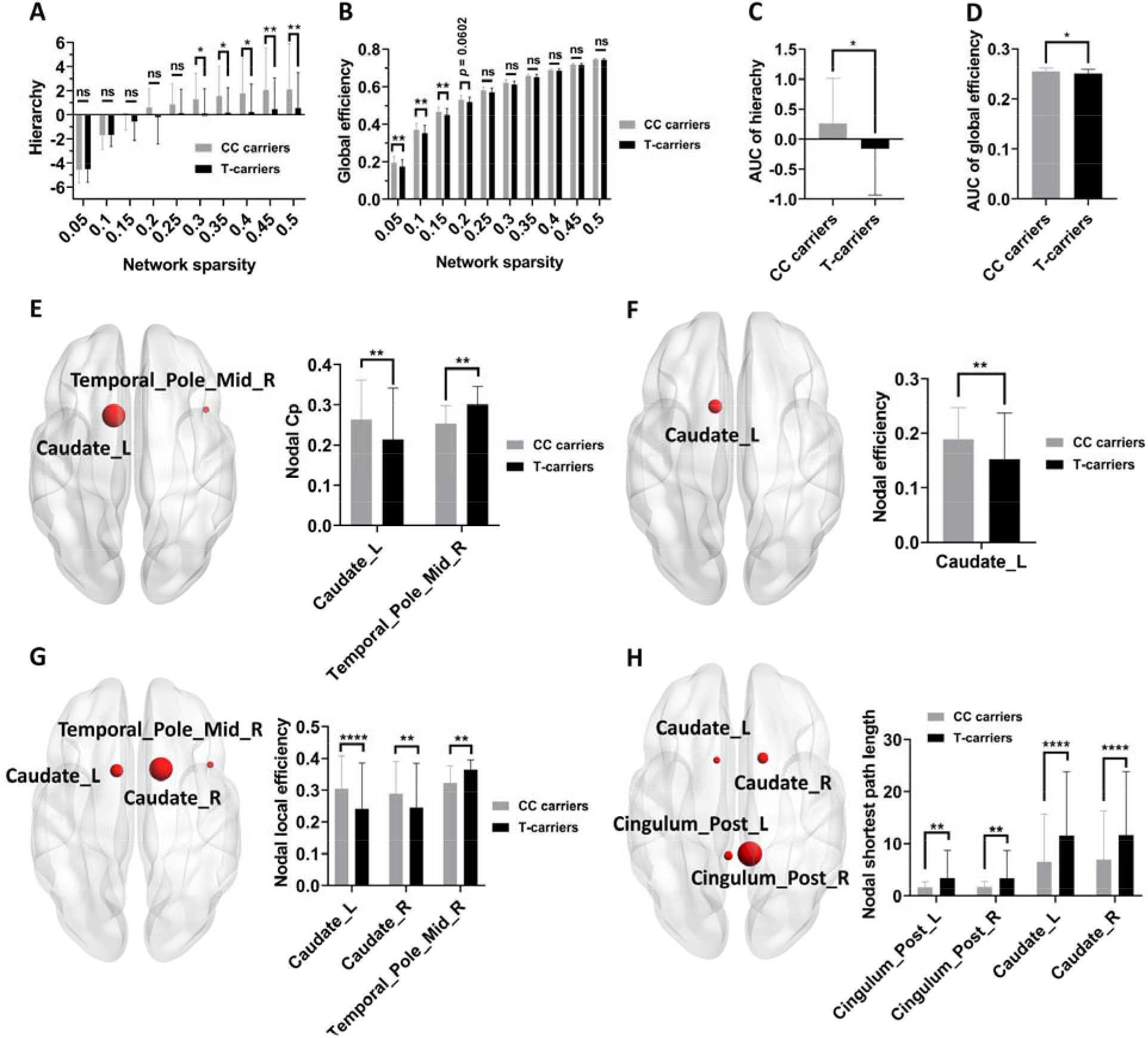
Group differences of functional network metrics between CC carriers and T-carriers. (A-B) Group differences of hierarchy (A) and global efficiency (B) at multiple network sparsity. (C-D) Group differences in the AUCs of hierarchy (C) and global efficiency (D). (E-H) Group differences of nodal Cp (E), nodal efficiency (F), nodal local efficiency (G), and nodal shortest path length (H). Two-way ANOVA test followed by FDR corrections was used for the analysis of results in A-B and E-H (*p* < 0.05 after FDR correction was considered statistically significant). The AUCs of global network metrics between CC carriers and T-carriers in C-D were compared using unpaired t-test (*p* < 0.05 was considered statistically significant). \**p* < 0.05, \*\**p* < 0.01, \*\*\*\**p* < 0.0001. Abbreviations: Cp, Clustering coefficient; AUC, Area under curve.

The global network metrics of functional network were not statistically different among CC carriers, CT carriers, and TT carriers (FDR-corrected *p* > 0.05). The TT carriers showed higher betweenness centrality in left precuneus and right paracentral lobule and lower betweenness centrality in right inferior temporal gyrus compared to CC and CT carriers (FDR-corrected *p* < 0.05; Fig.6A). The TT carriers showed higher degree centrality in bilateral amygdala, left putamen, left pallidum, left thalamus and lower degree centrality in bilateral inferior temporal gyrus, left fusiform, and left parietal gyrus compared to CT and TT carriers (FDR-corrected *p* < 0.05; Fig.6B). The nodal Cp, nodal efficiency, and nodal local efficiency of bilateral caudate were much lower in CT carriers but not in TT carriers compared to CC carriers (FDR-corrected *p* < 0.05; Fig.6C-F), except that the nodal local efficiency of left caudate was lower in TT carriers compared to CC carriers (FDR-corrected *p* < 0.05; Fig.6E). Similarly, we revealed higher nodal shortest path length of bilateral post cingulate gyrus in CT and TT carriers (FDR-corrected *p* < 0.05; Fig.6F).

**Fig. 6.**
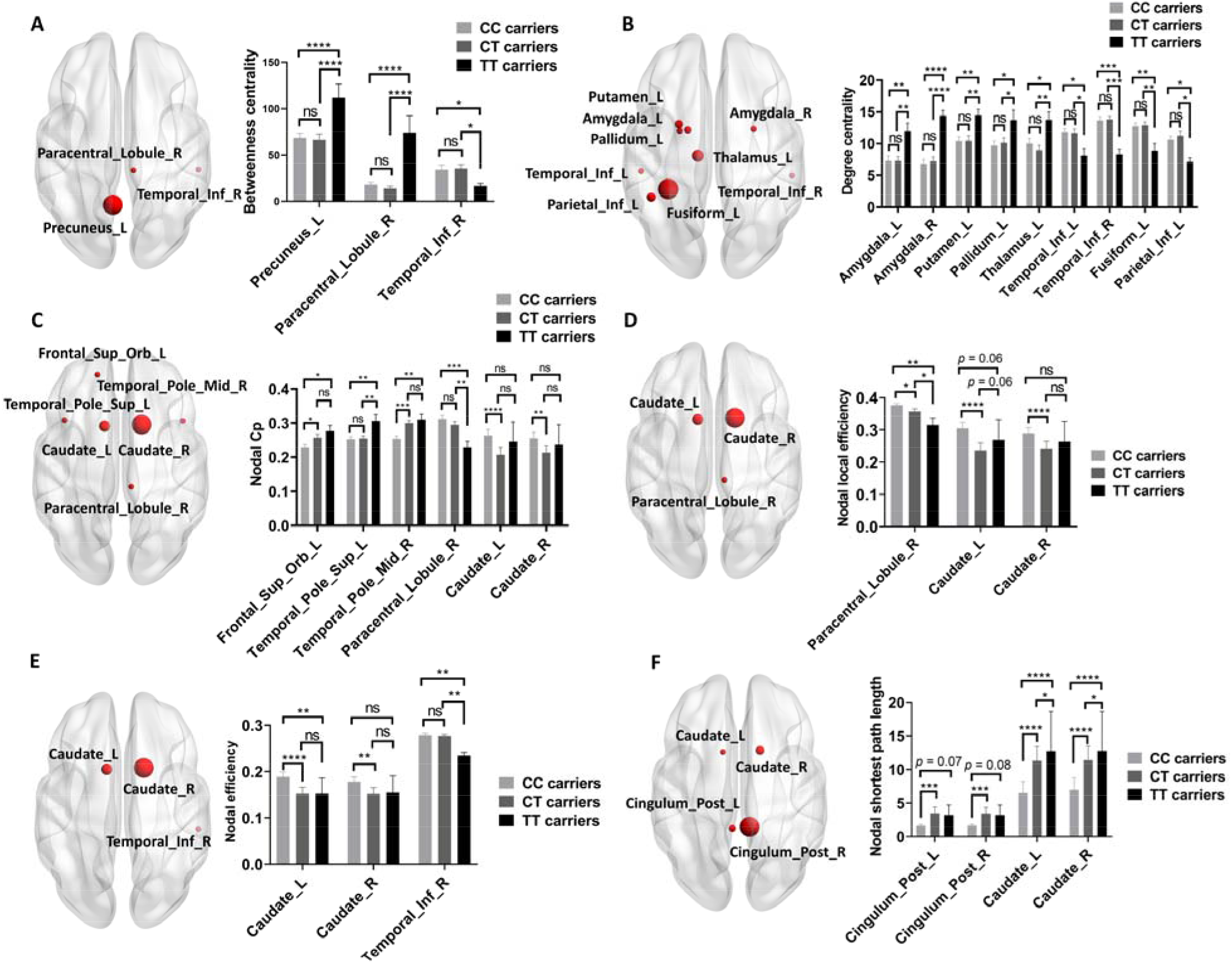
Group differences of functional network metrics among CC carriers, CT carriers, and TT carriers. (A-F) Group differences of nodal betweenness centrality (A), nodal degree centrality (B), nodal Cp (C), nodal efficiency (D), nodal local efficiency (E), and nodal shortest path length (F) in functional network. Two-way ANOVA test followed by FDR corrections was used and *p* < 0.05 after FDR correction was considered statistically significant. \**p* < 0.05, \*\**p* < 0.01, \*\*\**p* < 0.001, \*\*\*\**p* < 0.0001. Abbreviations: Cp, Clustering coefficient.

### The associations between *BIN3* rs2280104 and graphical networks

To examine whether the impacts of *BIN3* rs2280104 on graphical metrics of both structural and functional networks were independent of confounding variables, multivariate regression analysis was utilized to investigate the associations between *BIN3* rs2280104 and graphical metrics with age, sex, disease duration, and years of education as covariates. Table S2 showed that *BIN3* rs2280104 was significantly associated with multiple graphical metrics of structural network that were compared in Fig.3 and Fig.4. Table S3 shows that *BIN3* rs2280104 was significantly associated with multiple graphical metrics of functional network that were compared in Fig.5 and Fig.6.

### The associations between network metrics and ESS scores

The nodal Cp of left superior temporal pole in functional network was negatively associated with ESS scores (Pearson correlation analysis: r = –0.26, *p* = 0.0245 and Multivariate regression analysis: β = –26.79, *p* = 0.0156; Fig. 7A). The degree centrality of left calcarine in structural network was negatively associated with ESS scores (Pearson correlation analysis: r = –0.27, *p* = 0.001 and Multivariate regression analysis: β = –0.69, *p* = 0.0004; Fig. 7B).

**Fig. 7.**
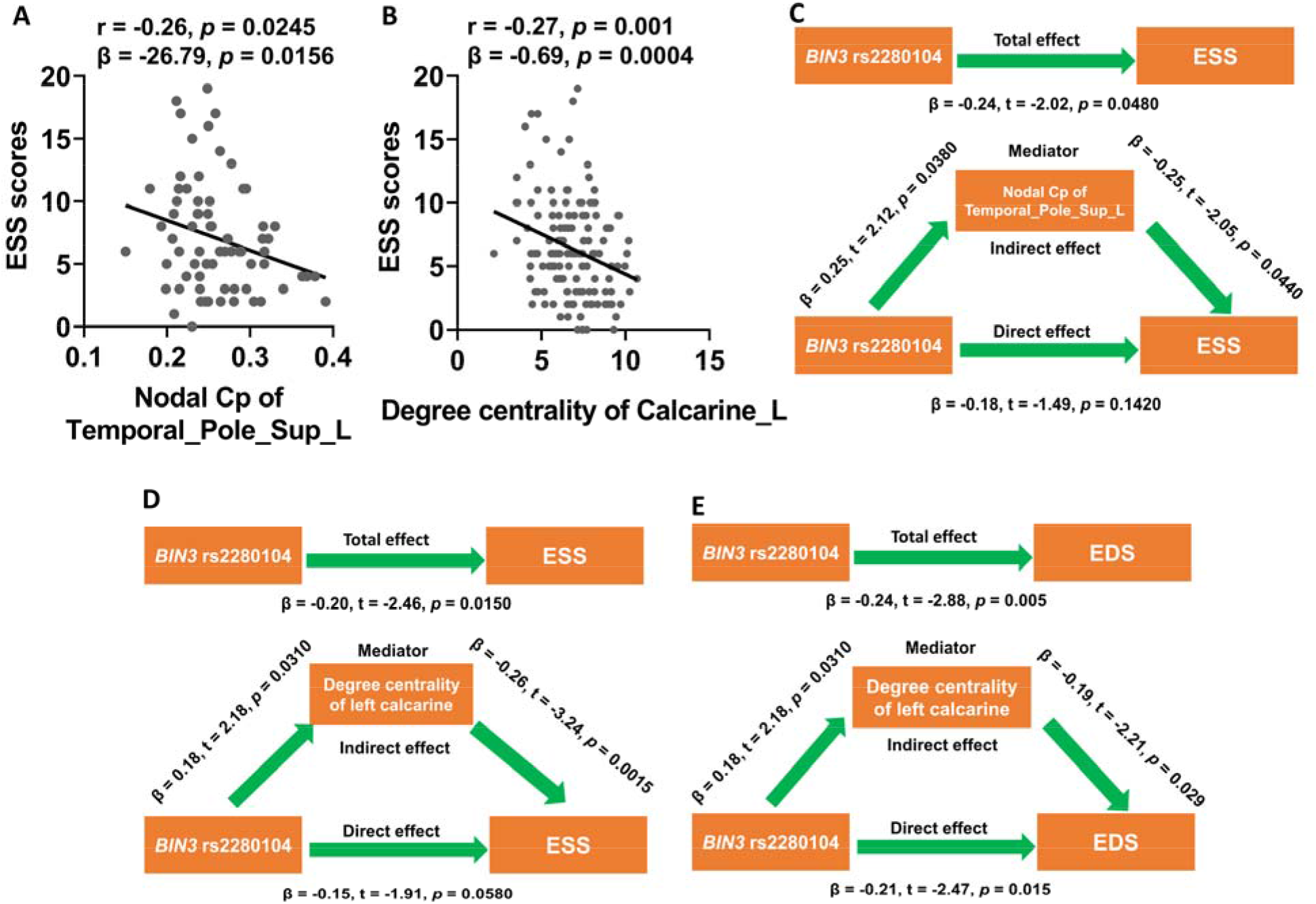
Mediation analysis suggested that the degree centrality of left calcarine in structural network mediated the effects of *BIN3* rs2280104 on EDS. (A-B) The nodal Cp of left superior temporal pole in functional network (A) and the degree centrality of left calcarine in structural network (B) were negatively associated with ESS scores (*p* < 0.05 in both Pearson correlation analysis and multivariate regression analysis). (C-D) The nodal Cp of left superior temporal pole in functional network (C) and the degree centrality of left calcarine in structural network (D) mediated the effects of *BIN3* rs2280104 on ESS scores. (E) The degree centrality of left calcarine in structural network mediated the effects of *BIN3* rs2280104 on EDS. The association analysis between network metrics and ESS scores was conducted by Pearson correlation method and multivariate regression analysis with age, sex, disease duration, and years of education as covariates. During the mediation analysis, age, sex, disease duration, and years of education were included as covariates. *p* < 0.05 was considered statistically significant. Abbreviations: EDS, Excessive daytime sleepiness; ESS, Excessive daytime sleepiness.

### Mediation analysis

The mediation analysis revealed that the nodal Cp of left superior temporal pole in functional network mediated the effects of *BIN3* rs2280104 on ESS scores (Fig. 7C). In addition, the degree centrality of left calcarine in structural network also mediated the effects of *BIN3* rs2280104 on ESS scores (Fig. 7D). Furthermore, when EDS was entered as dependent variable, we found the degree centrality of left calcarine in structural network but not nodal Cp of left superior temporal pole in functional network mediated the effects of *BIN3* rs2280104 on EDS (Fig. 7E).

## Discussion

In this study, we found EDS was associated with worse motor and non-motor symptoms in PD patients. Additionally, we revealed that PD-associated risk variant, *BIN3* rs2280104 was significantly associated with EDS in PD patients. Using network analysis, we demonstrated that *BIN3* rs2280104 prominently modified the structural and functional network metrics of PD patients. Both structural and functional network metrics were correlated with ESS scores of PD patients, however, only the degree centrality of left calcarine in structural network mediated the negative association between *BIN3* rs2280104 T allele on EDS.

## The associations between EDS and clinical features

EDS is one of most common sleep disturbances in PD ^46–49^. In our study, we found EDS+ patients exhibited higher rigidity scores, UPDRS-III scores, RBDSQ scores, SCOPA-AUT scores, and lower SDMT scores and striatum SBRs compared to EDS-patients, which were consistent with the results reported by previous studies. According to previous literature, it is generally recognized that EDS is associated with rapid motor progression and severe motor manifestations ^46, 51, 55^. In addition, EDS is also linked with worse non-motor symptoms, including depressive symptoms ^47, 48, 72^, autonomic dysfunction ^48, 72^, cognitive decline ^73–75^, and sleep disorders ^55, 76^. Furthermore, EDS is associated with increased frequency falls ^50, 77^, sarcopenia ^50^, development of swallowing impairment ^51^, and poorer quality of life (QoL) ^52, 53^. Essentially, EDS is also related to caudate denervation in PD ^75^. Taken together, previous findings and our results both support that EDS is an important clinical variable significantly associated with disease severity and progression in PD.

## The effects of *BIN3* rs2280104 T allele on EDS of PD patients

Although EDS is significantly associated with the clinical manifestations of PD patients, the neural mechanisms underlying EDS still remain unknown. Previous studies revealed several risk factors were potentially associated with EDS in PD, including age ^55, 57^, male sex ^55, 56^, worse sleep quality ^55–57^, depression ^58^, cognitive decline ^55, 56^, autonomic dysfunction ^58^, RBD ^55, 58^, and use of dopaminergic agents ^48, 56,57^ and antihypertensives ^56^. However, whether genetic factors contributed to EDS in PD was less understood. According to previous literature, only few studies investigated whether genetic variations were associated with EDS in PD patients. Recently, Adam et al. (2022) revealed that Human Leukocyte Antigen (*HLA*) risk allele DQB1*06:02 was significantly associated with EDS ^59^. *HLA-DQB1* gene is one of the members of *HLA* gene family and belongs to the major histocompatibility complex (MHC) class II beta chain paralogs. MHC class II molecule is a membrane-anchored heterodimer composed of an alpha (DQA) and a beta chain (DQB). It is responsible for the presentations of extracellular peptides from antigen-presenting cells to T or B cells in immune responses. *HLA-DQB1* gene was tightly associated with Narcolepsy type 1, which is a chronic sleep disorder caused by the loss of hypothalamic neurons producing wake-promoting hypocretin (also known as orexin) peptides ^78^. It has been shown that autoreactive CD4+ and CD8+ T cells specifically targeted self-antigen of hypocretin neurons and destructed these wake-promoting neurons in hypothalamus, which contributed to the sudden daytime sleepiness in Narcolepsy type 1 ^79, 80^. According to these results, it was possible that reduction of hypocretin neurons might mediate the association between *HLA*-*DQB1*\*06:02 and EDS in PD ^81, 82^. Indeed, Thannicka et al. (2007) have revealed loss of hypocretin neurons in PD and the percentage loss of hypocretin neurons gradually increased from Braak stage I (23%) to Braak stage V (62%) ^81^. Further study was required to examine whether *HLA*-*DQB1*\*06:02 was associated with the loss of hypocretin neurons in PD, which would help to validate above hypothesis. Another study also reported catechol-O-methyltransferase (*COMT*) polymorphism (rs4680) was associated with EDS in PD patients ^83^, however, the results were not confirmed in a large group of patients with PD ^84^. In our study, we found *BIN3* rs2280104 was significantly and negatively correlated with ESS scores in drug-naïve PD patients, which was independent of age, sex, education, and disease duration. In addition, based on our results, 20.75% of CC carriers (n = 53) had EDS, in contrast, only 6.45% of T-carriers (n = 93) had EDS. Interestingly, no EDS was found in TT-carriers (n = 21) of *BIN3* rs2280104. This finding was consistent with our recent study showing that *BIN3* rs2280104 was significantly associated with ESS scores in 198 PD patients from PPMI cohort ^62^. According to our study ^62^, the association between *BIN3* rs2280104 and ESS scores was also independent of other PD-associated risk variants assayed in PPMI ^62^. To summarize, our results demonstrated that *BIN3* rs2280104 was an essential genetic variant significantly associated with the heterogeneous distribution of EDS in PD patients. Future studies were required to explore whether other genetic variations were also associated with EDS in PD patients.

## The effects of *BIN3* rs2280104 T allele on structural network

Recently, we found that PD-associated risk variants significantly shaped structural and functional network metrics and brain network metrics mediated the effects of PD-associated risk variants on motor and non-motor features ^23, 62^. To understand the neural mechanisms underlying the effects of *BIN3* rs2280104 on EDS and disease susceptibility of PD patients, we investigated whether *BIN3* rs2280104 shapes graphical network metrics of structural network. For global network metrics, we found T-carriers showed higher global efficiency compared to CC carriers, which is consistent with lower small-worldness Lp in T-carriers. In fact, the changes of global efficiency and small-worldness Lp in structural network have been revealed in a recent meta-analysis ^85^. However, there were no significant differences in global network metrics among CC carriers, CT carriers, and TT carriers. These results indicated that the doses of T alleles didn’t significantly affect the effects of *BIN3* rs2280104 on the global network metrics of structural network. For nodal network metrics, we found reduced betweenness centrality in left middle occipital gyrus and left putamen in T-carriers compared to CC-carriers. In addition, both TT carriers and CT carriers exhibited decline of betweenness centrality in left middle occipital gyrus and left putamen. However, T allele dose-dependent reduction of betweenness centrality in left middle occipital gyrus and left putamen were not found in TT carriers and CT carriers. In our recent study, we revealed a PD-associated risk variant, *MAPT* rs17649553 significantly modified betweenness centrality of left putamen ^23^, which indicated that left putamen was a shared target of PD-associated risk variants. Because betweenness centrality in both left putamen and right putamen were modified by *BIN3* rs2280104, it was possible that basal ganglia network in structural network was specifically modified by *BIN3* rs2280104. In addition to basal ganglia network, the network metrics of some key nodes in visual network were also shaped by *BIN3* rs2280104, such as left middle occipital gyrus, left calcarine, left cuneus, left superior occipital gyrus, and left inferior occipital gyrus. These results supported that *BIN3* rs2280104 specifically shaped the key nodes in basal ganglia network and visual network, which were found to be disrupted in PD ^86–88^.

## The effects of *BIN3* rs2280104 T allele on functional network

We also examined whether *BIN3* rs2280104 affected network metrics of functional network. At the global level, we found reduced hierarchy and global efficiency in T-carriers compared to CC carriers. Hierarchy has been recognized as a key principle underlying the organization of human brain networks ^89^. The reduction of hierarchy in T-carriers indicated that *BIN3* rs2280104 T allele disrupted the hierarchical organizations of functional networks in PD. The global efficiency represents the information efficiency of networks at the global level. The reduced global efficiency of functional network has been reported in PD patients ^90, 91^. The reduced global efficiency in T-carriers suggested that the information efficiency of functional network was much lower in T-carriers than CC carriers. Similar to structural network, we found no dose-dependent changes of global network metrics in functional network among CC carriers, CT carriers, and TT carriers. We found lower nodal Cp, efficiency, local efficiency, and higher shortest path length of left caudate in T-carriers, indicating that left caudate in basal ganglia network was specifically targeted by *BIN3* rs2280104. This notion was further demonstrated by our results showing that reduced nodal Cp, efficiency, local efficiency, and higher shortest path length of bilateral caudate in CT carriers, with encouraging trend in TT carriers. These results were consistent with the findings reported by previous studies showing that nodal topology of caudate in functional network were significantly altered in PD ^92–95^. The reduction of local information efficiency in caudate of T-carriers indicated that *BIN3* rs2280104 T allele is harmful in PD, which was consistent with previous study showing that *BIN3* rs2280104 T allele was associated with increased risk of PD. We found TT carriers had no significant differences in multiple nodal network metrics of bilateral caudate, which may be due to the small sample size of TT carriers (n = 7) in functional network analysis. Future studies were required to examine whether the doses of T alleles in *BIN3* rs2280104 affected the local network topology of bilateral caudate in a larger population of PD patients.

## The associations between graphical network metrics and EDS

According to previous literature, both graphical network metrics in functional network and structural network were found to be correlated with the clinical manifestations of PD patients ^7, 23, 91^. Thus, we analyzed whether graphical network metrics modified by *BIN3* rs2280104 were correlated with ESS scores. As mentioned above, we found the nodal Cp of left superior temporal pole in functional network was negatively associated with ESS scores of PD patients. In fact, a previous study has revealed that the functional connectivity of left superior temporal pole has been revealed to be associated with sleep quality in healthy individuals ^96^. We found the degree centrality of left calcarine in structural network was negatively associated with ESS scores, while it is still unknown how left calcarine affect EDS of PD patients. Left calcarine is one of the key nodes in visual system. Several studies have reported structural and functional metrics in left calcarine were significantly altered in patients with sleep disturbances. For example, the functional connectivity between locus coeruleus and left calcarine was significantly enhanced in chronic insomnia disorder ^97^. In PD patients with RBD, the functional connectivity between right superior occipital gyrus and left calcarine was significantly reduced compared to patients without RBD ^98^. poor sleepers in patients with cerebral small vessel disease (CSVD) exhibited decreased gray matter volume in the left calcarine cortex compared to good sleepers of CSVD patients ^99^. In addition, gray matter volume in left calcarine cortex was associated with sleep quality in CSVD patients ^99^. Although both nodal Cp of left superior temporal pole in functional network and degree centrality of left calcarine in structural network were negatively associated with ESS scores, it was the degree centrality of left calcarine in structural network mediated the effects of *BIN3* rs2280104 on both ESS scores and EDS. Future studies were required to dissect the role of left calcarine in EDS of PD patients.

## The potential molecular mechanisms associated with the effects of *BIN3* rs2280104

According to GTEx database, *BIN3* rs2280104 was significantly associated with the changes of expressions of multiple genes (https://www.gtexportal.org/home/). Specifically, *BIN3* rs2280104 T allele was correlated with the reduced expressions of *BIN3* and *C8orf58* in a multitude of brain regions compared to C allele ^62^. In contrast, the expression of *PDLIM2* was significantly increased in T-carriers compared to CC carriers ^62^. *BIN3* gene encodes bridging integrator-3, which is a BAR domain adapter protein involved in the regulation of endocytosis, cell motility, carcinogenesis, and myogenesis ^100–104^. It has been shown that loss of *BIN3* resulted in cataracts and higher susceptibility to lymphoma in a mouse model ^101^. Bankston et al. (2013) revealed that During early myogenesis, BIN3 enhanced the migration of differentiated muscle cells by colocalizing with F-actin in lamellipodia ^105^. In addition, BIN3 specifically regulated the activity of Rho GTPases Rac1 and Cdc42 to promote myotube formation in differentiated muscle cells^105^. BIN3 was thought to be a tumor-suppressor in esophagus carcinoma, which is significantly related to the infiltration level of regular T cells, Treg cells, B cells, NK cells, and M2 macrophages. In contrast, the role of BIN3 in the pathogenesis of PD remained elusive. It has been reported that pesticides regulated the expressions of *BIN3* to increase PD risk ^104^, nevertheless, whether BIN3 mediated the effects of pesticides on PD was unknown. Previous studies revealed PD-associated risk variants were associated with common pathological pathways, including endocytosis, autophagy, lysosome, mitochondria metabolism, DNA replication, synaptic vesicle recycling, and microtubule polymerization ^60, 106, 107^, whether and how BIN3 participated in these biological processes were also unknown and deserved to be further explored in future studies.

## Strengths and limitations of this study

In this study, we duplicated previous findings that EDS was an essential non-motor symptom significantly associated with worse disease severity, which suggested that EDS should be carefully considered during the clinical management of PD. Using multivariate regression analysis and χ^2^ test, we demonstrated that a PD-associated risk variant, *BIN3* rs2280104 was specifically associated with EDS of PD patients. In addition to its effects on EDS and disease susceptibility, *BIN3* rs2280104 dramatically shaped the network topology of structural and functional network. Importantly, we found both the nodal Cp of left superior temporal pole in functional network and the degree centrality of left calcarine in structural network were negatively associated with ESS scores, which indicated that graphical network metrics were potential predictors of EDS in PD patients. Interestingly, with mediation analysis, we demonstrated that the degree centrality of left calcarine in structural network mediated the effects of *BIN3* rs2280104 on EDS, which provided network-based interpretations for effects of *BIN3* rs2280104 on EDS in PD. One of the limitations in this study was the lack of consideration of comorbidities, such as obstructive sleep apnea-hypopnea syndrome (OSAHS), which was also associated with daytime sleepiness in PD. Future studies were required to evaluate whether OSAHS contributed to EDS in PD patients. Another limitation was that we didn’t investigate the molecular mechanisms underlying the effects of *BIN3* rs2280104 on EDS in PD, further studies were encouraged to resolve this issue.

## Conclusions

In this study, we found *BIN3* rs2280104 is significantly associated with EDS of PD patients. Additionally, we also revealed that *BIN3* rs2280104 dramatically shaped the structural and functional network topology of PD patients. Importantly, we demonstrated that it was the degree centrality of left calcarine in structural network mediated the effects of *BIN3* rs2280104 on EDS. Future researches were required to identify the molecular mechanisms underlying the effects of *BIN3* rs2280104 on EDS and brain network metrics of PD patients.

## Author contributions

Zhichun Chen, Conceptualization, Formal analysis, Visualization, Methodology, Writing – original draft, Writing – review and editing; Bin Wu, Validation, Investigation, Methodology; Guanglu Li, Data curation, Formal analysis, Visualization; Liche Zhou, Data curation, Formal analysis, Investigation; Lina Zhang, Formal analysis, Investigation, Methodology; Jun Liu, Conceptualization, Supervision, Funding acquisition, Writing – original draft, Project administration, Writing-review and editing.

## Acknowledgments

Data used in the preparation of this article were obtained from the Parkinson’s Progression Markers Initiative (PPMI) database (www.ppmiinfo.org/data). We thank the share of PPMI data by all the PPMI study investigators. PPMI – a public-private partnership – is funded by the Michael J. Fox Foundation for Parkinson’s Research and funding partners, which can be found at www.ppmiinfo.org/fundingpartners.

## Funding information

This work was supported by grants from National Natural Science Foundation of China (Grant No. 81873778, 82071415) and National Research Center for Translational Medicine at Shanghai, Ruijin Hospital, Shanghai Jiao Tong University School of Medicine (Grant No. NRCTM(SH)-2021-03).

## Conflict of Interest

The authors have no conflict of interest to report.

## Data availability

All the raw data used in the preparation of this Article were downloaded from PPMI database (www.ppmi-info.org/data). All data produced in the present study are available upon reasonable request to the authors.

## Supporting Information

Additional supporting information may be found online in the Supporting Information section at the end of the article.

## Notes

### Competing Interest Statement

The authors have declared no competing interest.

