## Supplemental file for "*BIN3* rs2280104 T allele is associated with excessive daytime sleepiness and altered network topology in Parkinson’s disease"

**Supplementary materials**

**Table S1** The comparisons of demographic variables between EDS+ and EDS- patients

| Characteristic | EDS- patients  (n = 127) | EDS+ patients  (n = 17) | Statistic *p*-values | |
| --- | --- | --- | --- | --- |
| Age (years) | 61.06 ± 9.28 | 62.34 ± 9.16 |  | *p* > 0.05 |
| Sex (Female/Male) | 49/78 | 4/13 |  | *p* > 0.05 |
| Education (years) | 15.36 ± 2.99 | 14.82 ± 2.90 |  | *p* > 0.05 |
| Disease duration (years) | 2.08 ± 2.27 | 1.88 ± 0.96 |  | *p* > 0.05 |
| Age at onset (years) | 58.99 ± 9.47 | 60.46 ± 9.10 |  | *p* > 0.05 |

Values are shown as mean ± standard deviation. Sex ratio was evaluated with Chi-square test, other characteristics were compared using unpaired t-test. Abbreviations: EDS, Excessive daytime sleepiness.

**Table S2** The associations between ESS scores and clinical assessments

| **Motor assessments** | | | | | | | | | | | | | | | | | |
| --- | --- | --- | --- | --- | --- | --- | --- | --- | --- | --- | --- | --- | --- | --- | --- | --- | --- |
| HY | | | | Tremor | | | | | Rigidity | | | | | UPDRS-III | | | |
| β = 0.02  *p* = 0.0508 | | | | β = -0.06  *p* = 0.3680 | | | | | β = 0.15  *p* = 0.0192 | | | | | β = 0.60  *p* = 0.0042 | | | |
| **Non-motor assessments** | | | | | | | | | | | | | | | | | |
| RBDSQ | | SCOPA-AUT | | | LNS | | BJLOT | | | | SFT | | SDMT | | | MoCA | |
| β = 0.27  *p* < 0.0001 | | β = 0.59  *p* < 0.0001 | | | β = -0.10  *p* = 0.0639 | | β = -0.11  *p* = 0.0063 | | | | β = -0.21  *p* = 0.3817 | | β = -0.71  *p* = 0.0006 | | | β = -0.18  *p* = 0.0013 | |
| **Striatum SBR (n = 141/146)** | | | | | | | | | | | | | | | | | |
| Caudate_R | Caudate_L | | Putamen_R | | | Putamen_L | | Striatum_R | | Striatum_L | | Bilateral caudate | | | Bilateral putamen | | Bilateral striatum |
| β = -0.03  *p* = 0.0114 | β = -0.03  *p* = 0.0106 | | β = -0.01  *p* = 0.0497 | | | β = -0.01  *p* = 0.0611 | | β = -0.04  *p* = 0.0132 | | β = -0.04  *p* = 0.0142 | | β = -0.03  *p* = 0.0060 | | | β = -0.01  *p* = 0.0243 | | β = -0.02  *p* = 0.0064 |

The data were shown as the β and *p* values derived from multivariate regression analysis with age, sex, years of education, and disease duration as covariates. The motor function examination was assessed in ON state. Abbreviations: HY, Hoehn & Yahr stage; UPDRS-III, Unified Parkinson’ s Disease Rating Scale Part III; RBDSQ, REM Sleep Behavior Disorder Screening Questionnaire; SCOPA-AUT, Scale for Outcomes in Parkinson's Disease-Autonomic; SDMT, Symbol Digit Modalities Test; LNS, Letter Number Sequencing; SFT, Semantic Fluency Test Score; BJLOT, Benton Judgement of Line Orientation; MoCA, Montreal Cognitive Assessment; SBR, striatal binding ratio.

**Table S3** The associations between *BIN3* rs2280104 and graphical metrics of structural network

| Global efficiency | | | | | | | | | | | | | | |
| --- | --- | --- | --- | --- | --- | --- | --- | --- | --- | --- | --- | --- | --- | --- |
| Sparsity=0.15 | Sparsity=0.2 | | | Sparsity=0.25 | | | Sparsity=0.3 | Sparsity=0.35 | Sparsity=0.4 | | | Sparsity=0.45 | | Sparsity=0.5 |
| β = 0.0076  *p* = 0.1408 | β = 0.0076  *p* = 0.0704 | | | β = 0.0076  **p =* 0.0469 | | | β = 0.0076  **p* = 0.0352 | β = 0.0076  **p* = 0.0282 | β = 0.0076  **p* = 0.0235 | | | β = 0.0076  **p* = 0.0201 | | β = 0.0076  **p =* 0.0176 |
| **Small-worldness Lp** | | | | | | | | | | | | | | |
| Sparsity=0.15 | Sparsity=0.2 | | | Sparsity=0.25 | | | Sparsity=0.3 | Sparsity=0.35 | Sparsity=0.4 | | | Sparsity=0.45 | | Sparsity=0.5 |
| β = -0.04  *p* = 0.1112 | β = -0.04  *p* = 0.0556 | | | β = -0.04  **p =* 0.0371 | | | β = -0.04  **p* = 0.0278 | β = -0.04  **p* = 0.0222 | β = -0.04  **p* = 0.0185 | | | β = -0.04  **p* = 0.0159 | | β = -0.04  **p =* 0.0139 |
| **Nodal betweenness centrality** | | | | | | | | **Nodal degree centrality** | | | | | | |
| Occipital_Mid_L | | Putamen_L | | | Putamen_R | | | Calcarine_L | | Cuneus_L | | | Occipital_Sup_L | |
| β = -16.55  **p* = 0.0267 | | β = -19.65  *p* = 0.0716 | | | β = -12.79  *p* = 0.2088 | | | β = 0.44  **p* = 0.0311 | | β = 0.48  **p* = 0.0156 | | | β = 0.48  **p* = 0.0408 | |
| **Nodal Cp** | | | | | | | | **Nodal local efficiency** | | | | | | |
| Amygdala_R | | | Occipital_Inf_L | | | Angular_L | | Frontal_Mid_Orb_R | | | Amygdala_R | | | |
| β = 0.0197  *p* = 0.1062 | | | β = 0.0220  **p* = 0.0459 | | | β = 0.0188  **p* = 0.0351 | | β = 0.0208  *p* = 0.1184 | | | β = 0.0214  *p=* 0.1060 | | | |

Multivariate regression analysis was performed by adjusting age, sex, disease duration, and years of education. False discovery rate (FDR) was used for multiple comparison corrections. FDR-corrected *p*-values were shown. FDR-corrected *p* < 0.05 was considered statistically significant. **p* < 0.05.

**Table S4** The associations between *BIN3* rs2280104 and graphical metrics of functional network

| Hierarchy | | | | | | | | | | | | | | | | | | | | | | | | | | |
| --- | --- | --- | --- | --- | --- | --- | --- | --- | --- | --- | --- | --- | --- | --- | --- | --- | --- | --- | --- | --- | --- | --- | --- | --- | --- | --- |
| Sparsity=0.3 | | | | Sparsity=0.35 | | | | | | | Sparsity=0.4 | | | | Sparsity=0.45 | | | | | | | Sparsity=0.5 | | | | |
| β = -1.1450  *p* = 0.0638 | | | | β = -1.2960  *p* = 0.0890 | | | | | | | β = -1.3930  **p* = 0.0463 | | | | β = -1.4440  *p* = 0.0606 | | | | | | | β = -1.4040  *p =* 0.0826 | | | | |
| **Global efficiency** | | | | | | | | | | | | | | | | | | | | | | | | | | |
| Sparsity=0.05 | | | | | | Sparsity=0.1 | | | | | | | Sparsity=0.15 | | | | | | | | Sparsity=0.2 | | | | | |
| β = -0.0218  **p* = 0.0330 | | | | | | β = -0.0225  *p* = 0.0584 | | | | | | | β = -0.0187  **p* = 0.0236 | | | | | | | | β = -0.0130  **p* = 0.0430 | | | | | |
| **Nodal betweenness centrality** | | | | | | | | | | | | **Nodal degree centrality** | | | | | | | | | | | | | | |
| Precuneus_L | | | Paracentral_Lobule_R | | | | | Temporal_Inf_R | | | | Amygdala_L | Amygdala_R | | | Putamen_L | | Pallidum_L | | | | | Thalamus_L | | Temporal_Inf_L | |
| β = 11.29  *p* = 0.1545 | β = 15.84  ***p* = 0.0030 | | | | | | β = -6.44  *p* = 0.1824 | | | | | β = 1.50  *p* = 0.1791 | β = 2.25  **p* = 0.0432 | | | β = 0.70  *p* = 0.4232 | | β = 0.80  *p* = 0.3883 | | | | | β = 0.20  *p* = 0.8084 | | β = -0.99  *p* = 0.2531 | |
| **Nodal degree centrality** | | | | | | | | | | | | **Nodal Cp** | | | | | | | | | | | | | | |
| Temporal_Inf_R | | | Fusiform_L | | | | | Parietal_Inf_L | | | | Frontal_Sup_Orb_L | | Temporal_Pole_Sup_L | | | Temporal_Pole_Mid_R | | | Paracentral_  Lobule_R | | | | Caudate_L | | Caudate_R |
| β = -1.63  *p* = 0.0527 | | | β = -0.99  *p* = 0.2516 | | | | | β = -0.72  *p* = 0.4305 | | | | β = 0.03  ***p* = 0.0081 | | β = 0.02  *p* = 0.0570 | | | β = 0.03  ****p* = 0.0006 | | | β = -0.03  **p* = 0.0104 | | | | β = -0.03  *p* = 0.2916 | | β = -0.02  *p* = 0.3224 |
| **Nodal efficiency** | | | | | | | | | **Nodal local efficiency** | | | | | | | **Nodal shortest path length** | | | | | | | | | | |
| Paracentral_Lobule_R | | Caudate_L | | | Caudate_R | | | | Caudate_L | Caudate_R | | | Temporal_Inf_R | | | Cingulum_Post_L | | | Cingulum_Post_R | | | | | Caudate_L | | Caudate_R |
| β = -0.03  ***p* = 0.0072 | | β = -0.04  *p* = 0.2102 | | | β = -0.03  *p* = 0.2933 | | | | β = -0.03  *p* = 0.1025 | β = -0.02  *p* = 0.2319 | | | β = -0.01  ***p* = 0.0045 | | | β = 1.36  *p* = 0.3340 | | | β = 1.29  *p* = 0.2028 | | | | | β = 3.41  *p* = 0.1519 | | β = 3.06  *p* = 0.1646 |

Multivariate regression analysis was performed by adjusting age, sex, disease duration, and years of education. FDR-corrected *p*-values were shown. FDR-corrected *p* < 0.05 was considered statistically significant. **p* < 0.05, ** *p* < 0.01, *** *p* < 0.001.

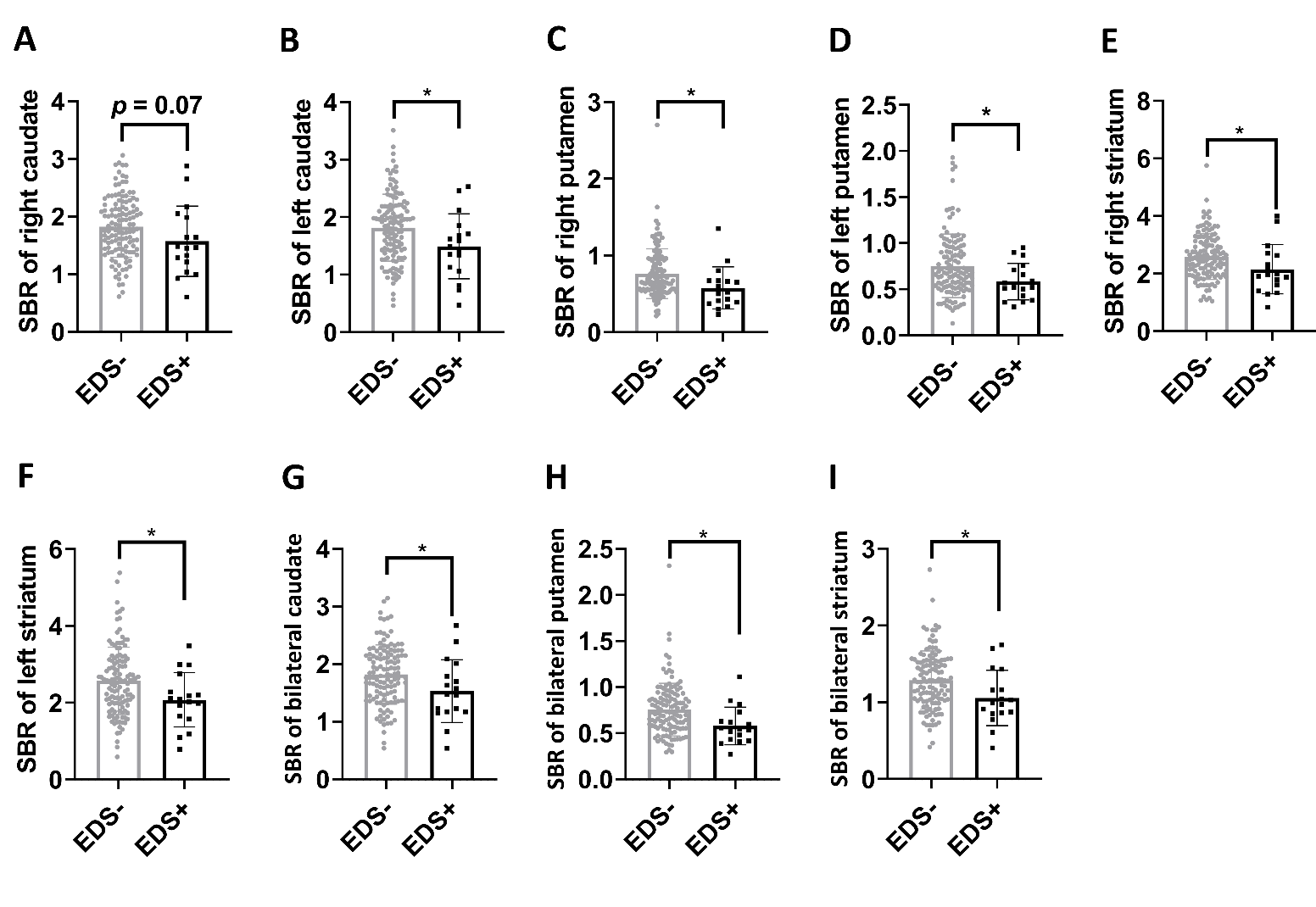

**Fig. S1 Group differences in the SBRs of striatum subregions between EDS- patients and EDS+ patients.** (A-I) Group differences of SBRs in right caudate (A), left caudate (B), right putamen (C), left putamen (D), right striatum (E), left striatum (F), bilateral caudate (G), bilateral putamen (H), and bilateral striatum (I). Unpaired t-test was used to compare the SBRs of striatum between EDS- patients and EDS+ patients. *p* < 0.05 was considered statistically significant. **p* < 0.05.
